# Deep learning-based approach for detecting signs of atrial septal defect on chest radiographs: a proof of concept study

**DOI:** 10.1101/2022.01.30.22270137

**Authors:** Ryo Matsuoka, Hiroshi Akazawa, Satoshi Kodera, Katsura Soma, Hiroki Yagi, Masahiko Umei, Hiroshi Kadowaki, Junichi Ishida, Hiroki Shinohara, Susumu Katsushika, Hirotaka Ieki, Toshihiro Yamaguchi, Yasutomi Higashikuni, Katsuhito Fujiu, Kaoru Ito, Atsushi Yao, Issei Komuro

## Abstract

Many patients with atrial septal defects (ASD) are asymptomatic and undiagnosed during the first few decades of life, but have overt heart failure, arrhythmias, cerebral infarction, and increased mortality in adults with advancing age. To provide a non-invasive, easy-to-use, and effective method for detecting ASD, we aimed to develop and validate a deep learning-based algorithm to diagnose ASD on chest radiographs. The ASD dataset was created from 173 chest radiographs of 74 patients with ASD and 170 chest radiographs of 100 patients without ASD. Convolutional neural network models (VGG16, ResNet50, DenseNet121, and Xception) for diagnosing ASD were pretrained using two different datasets, the large-scale real-world ImageNet dataset and the ChestX-ray14 dataset released by National Institutes of Health, followed by a round of training using the training set of the ASD dataset. Model performance was evaluated by five-fold stratified cross-validation. The best performance in ImageNet pretraining was achieved by ResNet50 model, and the cross-validation area under the curve (AUC) was 0.95, with sensitivity of 0.86, specificity of 0.87, and overall accuracy of 0.87. The best performance in ChestX-ray pretraining was achieved by Xception, and the cross-validation AUC was 0.93, with sensitivity of 0.85, specificity of 0.85, and overall accuracy of 0.85. The diagnostic performances of these models were comparable to those of cardiologists. Gradient-weighted Class Activation Mapping showed that the ImageNet-pretrained model focused on bilateral hilar regions, while the ChestX-ray14-pretrained model focused on areas around cardiac silhouette and lower lung fields. Our deep learning-based algorithms made a diagnosis of ASD on the input chest radiographs with high accuracy, and had potential to help clinicians make accurate diagnosis of ASD from routine chest radiography, leading to improvement of prognosis and quality of life in patients with ASD.

## Introduction

Atrial septal defect (ASD) is the most common among adult congenital heart diseases, when bicuspid aortic valve and mitral valve prolapse are excluded [1, 2], and accounts for 25-30% of congenital heart disease cases diagnosed in adulthood [3, 4]. Although most patients with isolated ASD remain asymptomatic throughout childhood, overt symptoms of fatigue, exercise intolerance, and shortness of breath become prevalent with advancing age, and patients with unrepaired ASD ultimately manifest heart failure leading to a shorter lifespan [3, 4]. Currently, surgical closure of isolated ASD is safe with a rare mortality, and less invasive transcatheter closure becomes an alternative treatment of choice for secundum type of ASD. Irrespectively of the procedure, closure of ASD in patients over 40 years of age provides benefits for morbidity and mortality [5, 6]. However, it was reported that, among patients with surgically repaired ASD, those operated on after the age of 25 had lower survival than healthy general population, although those operated on before the age of 25 had an excellent prognosis [7]. Manifestations of atrial fibrillation, heart failure, and stroke were significantly more frequent when surgical repair was performed in older patients [7]. In view of these clinical evidence, early diagnosis is crucial for ASD patients to undergo elective closure of the defect for improvement of their prognosis. However, in real-world practice, ASD is occasionally overlooked and undiagnosed for decades, because the symptoms can be subtle and nonspecific and technical expertise is required for clinicians to suspect ASD on routine physical examination, electrocardiography, and chest radiography.

Recent advances in artificial intelligence (AI) technology are dramatically transforming healthcare and medicine [8, 9]. Especially, image recognition by using deep learning is increasingly applied to identification, classification, and quantification of patterns in medical images [10], such as X-ray radiography [11, 12], echocardiography [13], computed tomography [14], and magnetic resonance imaging [15]. Deep learning is a subset of machine learning that employs multilayered neural networks to learn representations of the given input data with multiple levels of abstraction and provide the final output without help from humans [16, 17].

We hypothesized that a deep learning-based algorithm had potential to help clinicians to make accurate diagnosis of adult ASD. In this study, we built deep learning-based algorithms to detect signs of ASD from chest radiographs, and demonstrated that the performance of our models was comparable to that of cardiologists.

## Materials and Methods

### Patient recruitment and ASD dataset

We first recruited 99 patients with ASD who were aged 15 years or older and visited the Department of Cardiology at The University of Tokyo Hospital during the period from April 2005 to June 2020 (Fig 1). They were diagnosed to have ASD using transthoracic echocardiography, computed tomography, or magnetic resonance imaging. We excluded 25 patients; who had no chest radiographs before ASD closure; who had pacemaker implantation; who was diagnosed with Eisenmenger syndrome (Fig 1). We finally included 74 ASD patients, and collected their frontal-view and posterior-to-anterior chest radiographs taken prior to the procedures for ASD closure. For the non-ASD control group, we manually selected patients; who were confirmed not to have ASD using transthoracic echocardiography; whose age and sex were matched to ASD patients as closely as possible; who underwent coronary angiography or percutaneous coronary intervention at The University of Tokyo Hospital during the period from April 2005 to June 2020; who had no history of cardiac surgery; whose chest radiographs had no sign of acute heart failure. Their frontal-view and posterior-to-anterior chest radiographs were also collected (Fig 1). The chest radiographs were obtained using four different digital radiography systems (TOSHIBA MRAD-A80S/5G, FUJIFILM DR-ID600, GE Healthcare Revolution XR/d, SHIMADZU RADspeed Pro V4).

**Fig 1.**
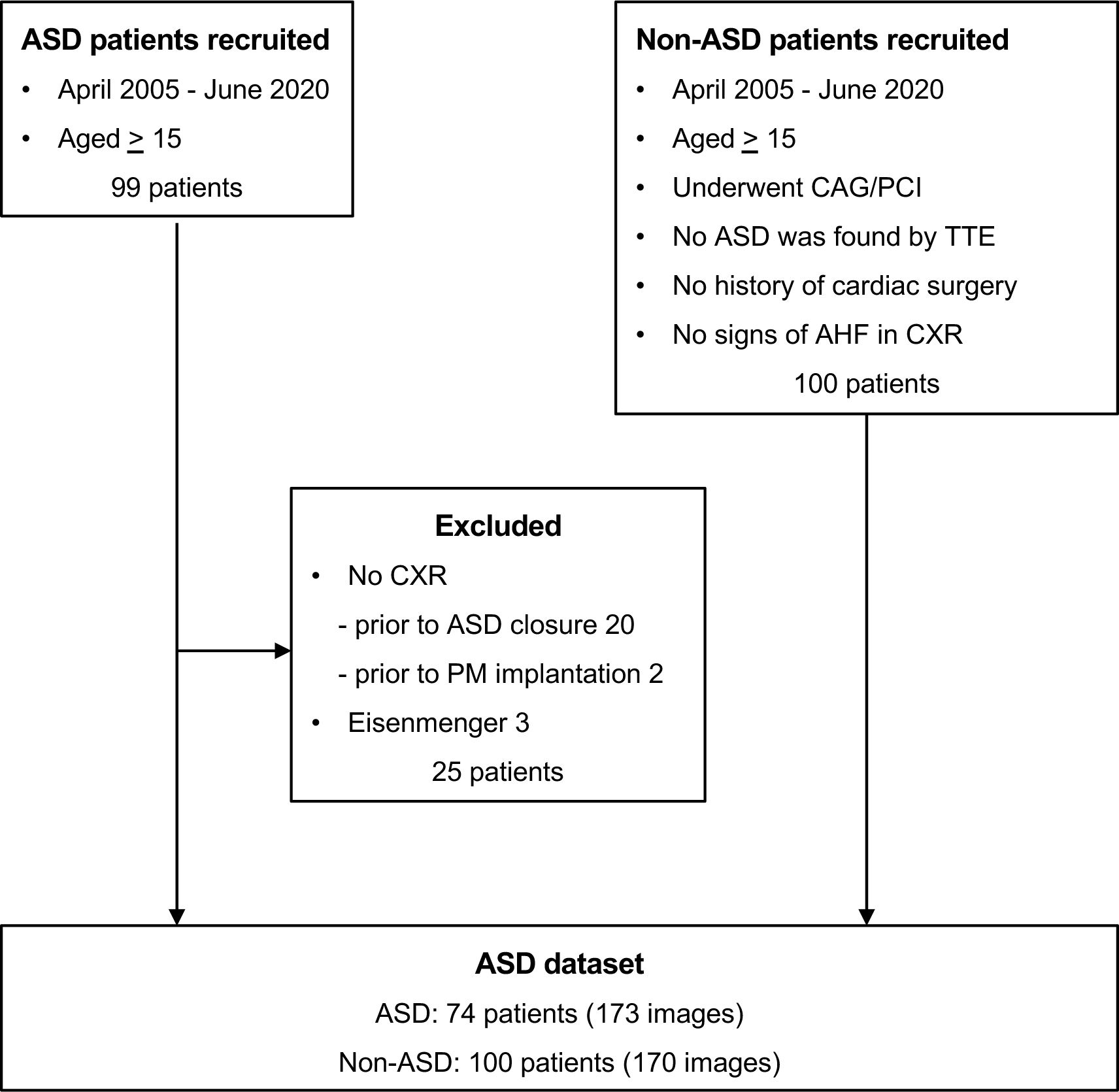
Selection of study population. AHF; acute heart failure, ASD; atrial septal defect, CAG; coronary angiography, CXR; chest X-ray, PCI; percutaneous coronary intervention, PM; pacemaker, TTE; transthoracic echocardiography.

We constructed “ASD dataset” by combining all these images (Fig 1). All images were manually cropped into a square, with the part outside the lung fields being removed, and then resized to 256 × 256 pixels. All procedures of this study were conducted under relevant guidelines and regulations and in compliance with the 1975 Declaration of Helsinki. This study was approved by the Institutional Review Board of the University of Tokyo (approval number 2650), and waiver of individual informed consent was granted by the institutional review board.

### Datasets for pretraining

To develop deep-learning predictive models, we adopted a transfer learning method, as previously described [18]. We first built convolutional neural networks (CNNs) pretrained on two different datasets, and then retrained them on our ASD dataset to compare the impacts of individual datasets for pretraining. The datasets we adopted for pretraining were the large-scale real-world ImageNet dataset and the ChestX-ray14 dataset released by National Institutes of Health. ImageNet dataset is a large collection of image data containing more than 1.2 million images of 1,000 object class for public use in visual object recognition research [19]. Chest radiographs were collected for pretraining from the ChestX-ray14 dataset, which contained 112,120 frontal-view chest radiographs of 30,805 unique patients [20]. We selected 1,563 posterior-to-anterior images of 1,150 patients whose finding labels contained the word “cardiomegaly”. In addition, we randomly picked up 10,000 posterior-to-anterior images of 7,545 patients whose finding labels did not contain the word “cardiomegaly”. All images were resized to 256 × 256 pixels. We defined these images as “ChestX-ray dataset”, and randomly split them into the training set : test set in an 8:2 ratio (Hold-out validation) (Fig 2).

**Fig 2.**
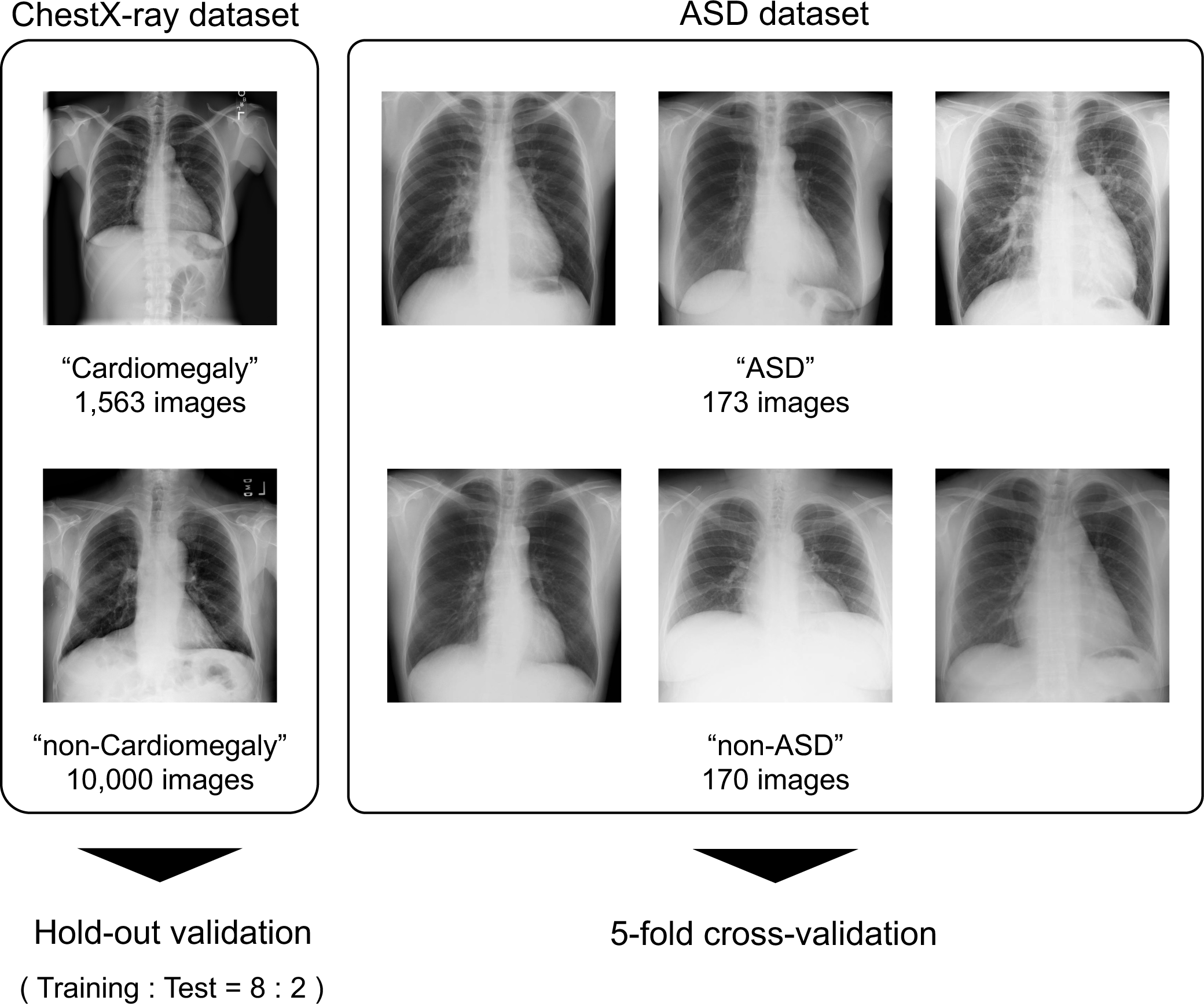
Data preparation. The “ChestX-ray dataset” consisted of 1,563 cardiomegaly images and 10,000 non-cardiomegaly images, collected from ChestX-ray 14 dataset. The “ASD dataset” consisted of 173 chest radiographs of ASD patients and 170 chest radiographs of non-ASD patients.

### Model architecture and pretraining

We used four state-of-the-art CNN architectures, VGG16 [21], ResNet50 [22], DenseNet121 [23], and Xception [24] as base model. We replaced the final layer with a global average pooling layer and a fully connected layer that had a single output, after which we applied a sigmoid nonlinearity for generating a predicted probability of ASD.

As described above, each base model was pretrained on two different datasets, ImageNet dataset and ChestX-ray dataset. The four models, VGG16, ResNet50, DenseNet121, and Xception, pretrained on the ImageNet dataset were available in the Keras library [25], and we used them as ImageNet-pretrained models, accordingly. For pretraining on ChestX-ray dataset, we used data augmentation technique: random rotation; random zooming; vertical and horizontal shifting; perspective distortion; random brightness.

### Model training and test

We retrained each of the two models on the ASD dataset, and assessed the detection performance using a stratified five-fold cross-validation approach [26]. Briefly, the entire ASD dataset was randomly split into five equal-sized subsets without overlapping of patients. After training on four subsets, we assessed performance of the model on the remaining subset. This process was repeated five times. We used all images per patient for training, but used one image per patient for test.

We plotted receiver operating characteristic (ROC) curves, and calculated the mean area under the curve (AUC). We selected the cut-off points from each training set using the Youden J statistics, and calculated mean sensitivity, specificity, and overall accuracy as comparative metrics. We used gradient-weighted class activation mapping (Grad-CAM) to visualize the areas of interest where the models focused [27].

### Interpretation by cardiologists

To ensure the validity of the test set and evaluate the performance of our models in diagnosing ASD from chest radiographs, we compared their performance with that of 3 physicians, all of whom were board-certified cardiologists with over 9 years clinical experience. They classified the images of the same test sets into ASD or non-ASD, and we calculated sensitivity, specificity, and overall accuracy of each cardiologist.

### Software

For data preparation, model training, and validation, we used Python 3.7 and its libraries including Tensorflow 1.15.2 [28] with graphical processing units, GeForce RTX 2070 SUPER (NVIDIA). Statistical analysis was performed using JMP Pro 15 (SAS Institute, Inc.).

## Results

### The ASD dataset

This study included 173 chest radiographs of 74 ASD patients (67 images of 27 male and 106 images of 47 female patients) (Table 1). The median age when chest radiographs were taken was 45.0 years. Among the ASD patients, 66 patients underwent right heart catheterization, and the average pulmonary-to-systemic flow ratio (Qp/Qs) was 2.19. Pulmonary hypertension was observed in 14 patients, and pulmonary vasodilators were administered in 11 patients (Table 1). This study also included 170 chest radiographs of 100 non-ASD patients who underwent coronary angiography or percutaneous coronary intervention (74 images of 42 male and 96 images of 58 female). The median age when chest radiographs were taken was 48.0 years.

**Table 1.** Characteristics of Patients.

|  | ASD | Non-ASD |
| --- | --- | --- |
| Patients, No. | 74 | 100 |
| Chest radiographs, No. | 173 | 170 |
| Male, No. of images (%) | 67 (39) | 74 (44) |
| Age when chest radiographs taken, median (IQR), years | 45.0 (33.0 - 60.0) | 48.0 (42.0-55.0) |
| Right heart catheterization, No. (%) | 66 (89) | - |
| Cardiothoracic Ratio, Mean (SD) | 46.2 (17.2) | 48.6 (10.1) |
| Pulmonary to systemic flow ratio, Mean (SD) | 2.19 (0.90) | - |
| Pulmonary hypertension |  |  |
| Mean PAP ≥ 25 mmHg, No. (%) | 14 (19) | - |
| Pulmonary vasodilators, No. (%) | 11 (15) | - |
| Type of ASD |  |  |
| Ostium secundum, No. (%) | 66 (89) | - |
| Sinus venosus, No. (%) | 3 (4) | - |
| Coronary sinus, No. (%) | 2 (3) | - |
| Mixed, No. (%) | 3 (4) | - |
| Clinical progress |  |  |
| Surgical closure, No. (%) | 16 (22) | - |
| Percutaneous closure, No. (%) | 37 (50) | - |
| Medication only, No. (%) | 4 (5) | - |
| Observation, No. (%) | 17 (23) | - |
| Indication for CAG |  |  |
| Acute coronary syndrome, No. (%) | - | 9 (9) |
| Angina Pectoris, No. (%) | - | 41 (41) |
| Follow-up CAG after PCI, No. (%) | - | 49 (49) |
| Takotsubo cardiomyopathy, No. (%) | - | 1 (1) |

CAG; coronary angiography, IQR; interquartile range, SD; standard deviation, PAP; pulmonary arterial pressure, PCI; percutaneous coronary intervention.

### Pretraining on the ChestX-ray14 dataset

Training set of the ChestX-ray dataset included 10,420 images of 7,857 patients, and test set included 1,143 images of 838 patients. We trained our untrained model on the training set, and assessed performance of the model for detecting cardiomegaly on the test set. The ResNet50-, DenseNet121-, and Xception-based models succeeded in learning and detected cardiomegaly with an AUC of 0.92, 0.91 and 0.93, respectively, but the VGG16-based model failed in learning even though we tried many parameters setting. Grad-CAM showed that our model made decision by looking at the cardiac shadow and area around the heart (S1Fig).

### Fine-tuning and model evaluation

We evaluated performance of the models using stratified five-fold cross validation. All models pretrained on the ImageNet dataset diagnosed ASD with an average AUC of around 0.90 (Table 2). The ResNet50-, DenseNet121-, and Xception-based models pretrained on the ChestX-ray dataset also diagnosed ASD with an average AUC of around 0.90 (Table 2). We selected the cut-off points, and calculated average sensitivity, specificity, and overall accuracy of each model (Table 2). The best performance in ImageNet pretraining was achieved by ResNet50 model, and the cross-validation AUC was 0.95, with sensitivity of 0.86, specificity of 0.87, and overall accuracy of 0.87. The best performance in ChestX-ray pretraining was achieved by Xception, and the cross-validation AUC was 0.93, with sensitivity of 0.85, specificity of 0.85, and overall accuracy of 0.85. The ROC curves of ImageNet-pretrained and ChestX-ray-pretrained models were shown in Fig3. We also plotted true positive rate and false positive rate of each cardiologist (Fig 3). The performances of the models were comparable or superior to those of cardiologists.

**Table 2.**
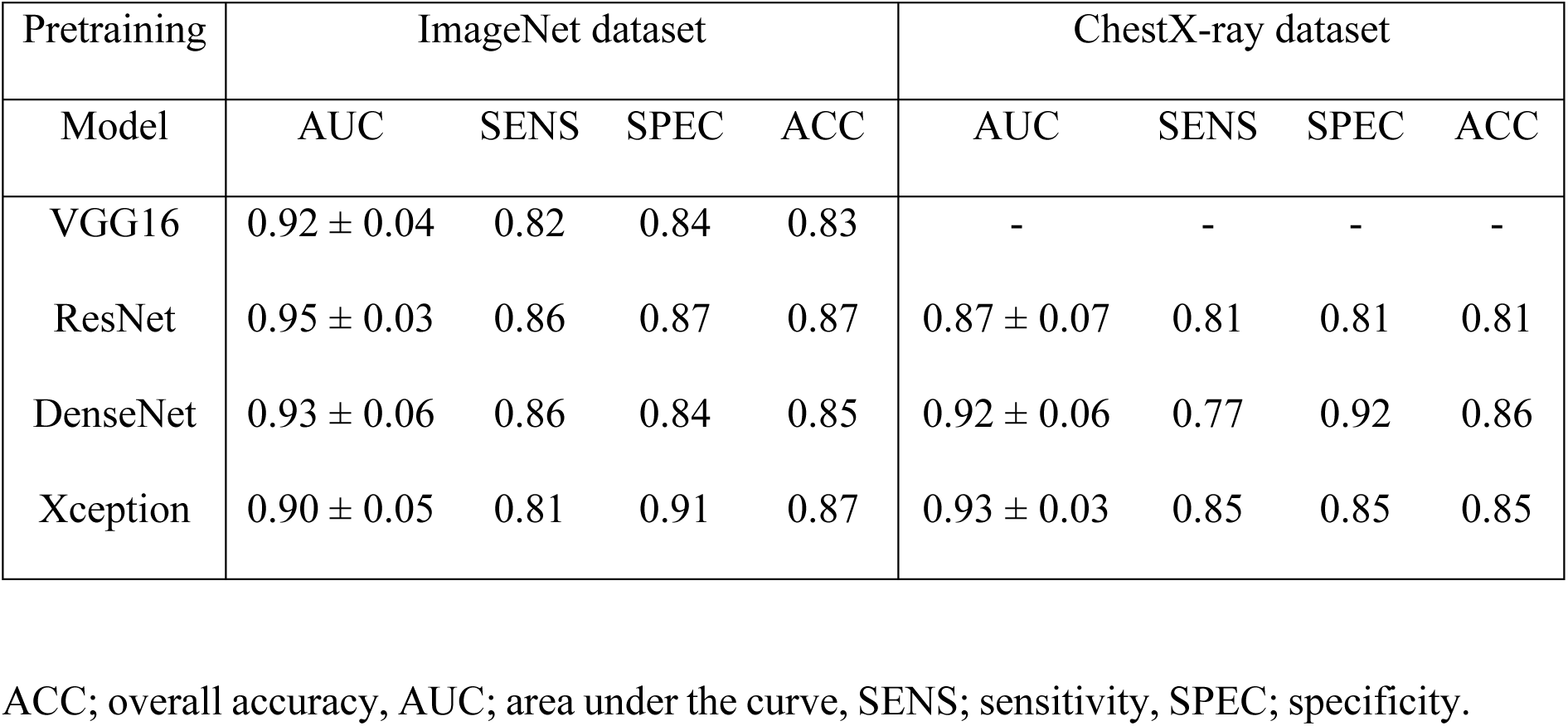
Performance metrics of the models.

| Pretraining | ImageNet dataset |  |  |  | ChestX-ray dataset |  |  |  |
| --- | --- | --- | --- | --- | --- | --- | --- | --- |
| Model | AUC | SENS | SPEC | ACC | AUC | SENS | SPEC | ACC |
| VGG16 | $0.92 \pm 0.04$ | 0.82 | 0.84 | 0.83 | - | - | - | - |
| ResNet | $0.95 \pm 0.03$ | 0.86 | 0.87 | 0.87 | $0.87 \pm 0.07$ | 0.81 | 0.81 | 0.81 |
| DenseNet | $0.93 \pm 0.06$ | 0.86 | 0.84 | 0.85 | $0.92 \pm 0.06$ | 0.77 | 0.92 | 0.86 |
| Xception | $0.90 \pm 0.05$ | 0.81 | 0.91 | 0.87 | $0.93 \pm 0.03$ | 0.85 | 0.85 | 0.85 |

ACC; overall accuracy, AUC; area under the curve, SENS; sensitivity, SPEC; specificity.

**Fig 3.**
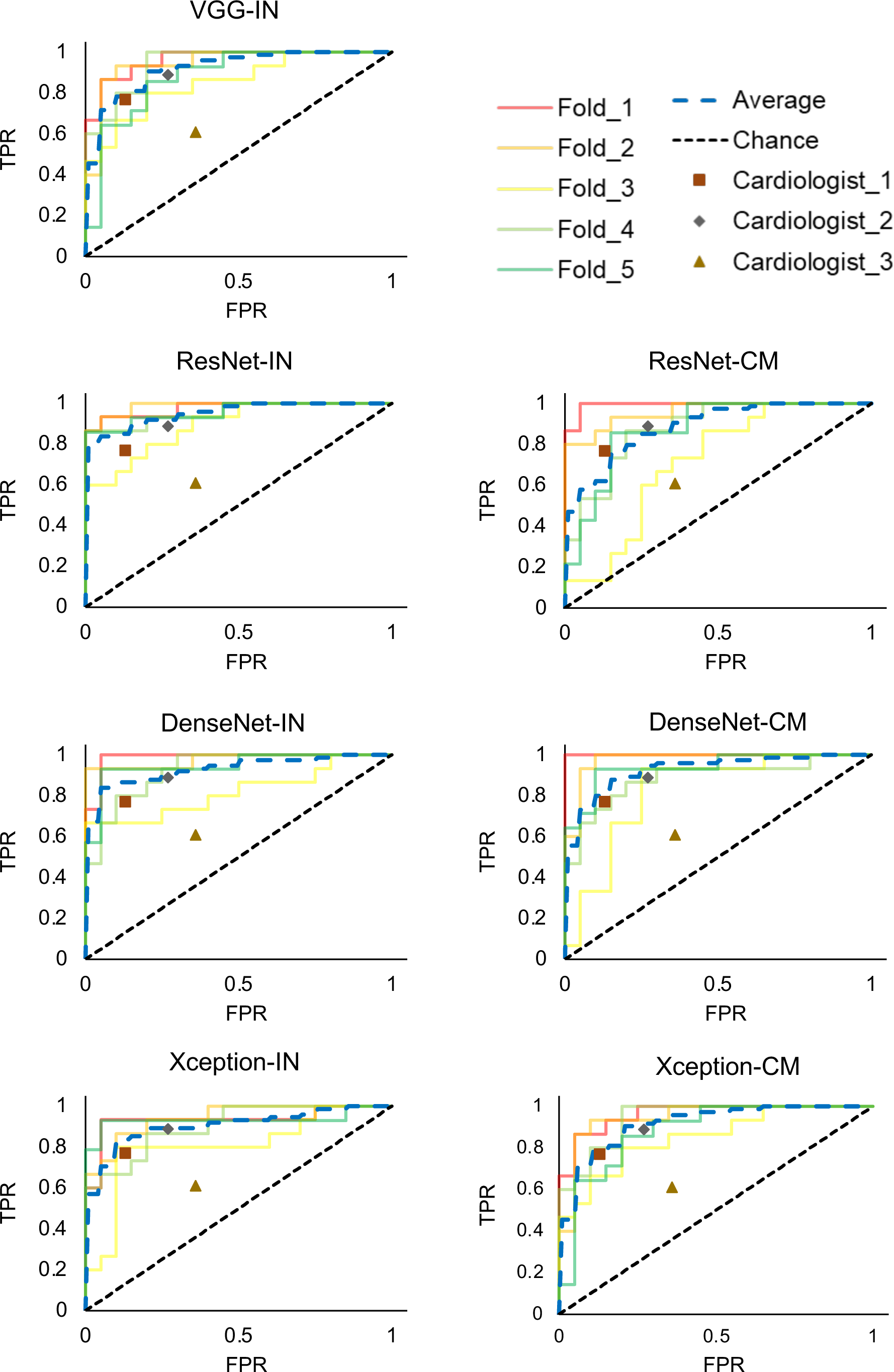
ROC curves of the models on the ASD dataset. ROC curves on every fold and average ROC curve were plotted. The true positive rates and false positive rates of three cardiologists were also plotted. CM; pretrained on the ChestX-ray dataset, FPR; false positive rate, IN; pretrained on the ImageNet dataset, TPR; true positive rate.

Grad-CAM showed that the areas where our models paid attention to interpret input images were different depending on the architecture and dataset for pretraining. The ImageNet-pretrained model focused on bilateral hilar regions, while ChestX-ray-pretrained model tended to focus on areas around cardiac silhouette and lower lung fields (Fig 4).

**Fig 4.**
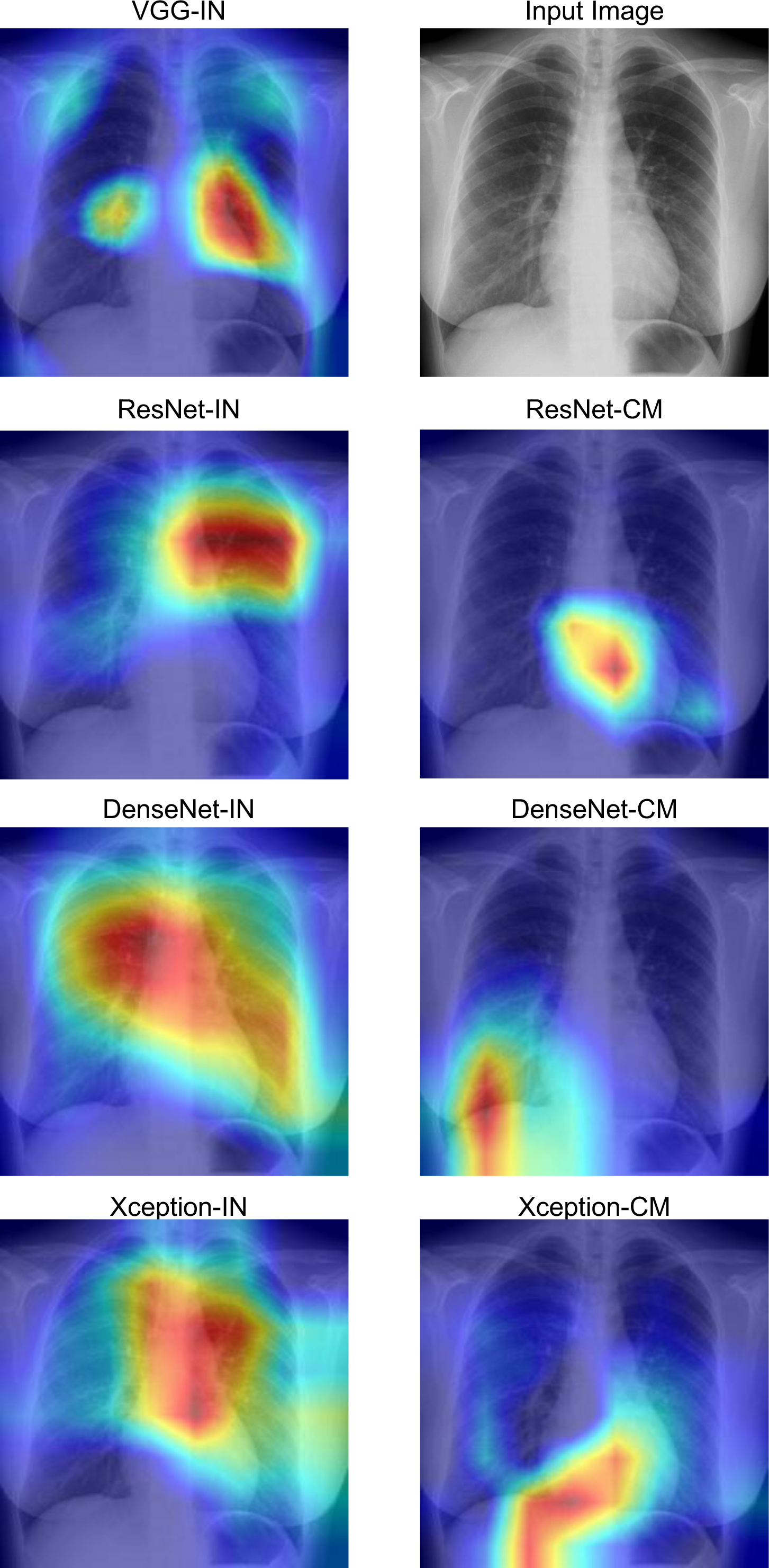
An example of Grad-CAM of a chest radiograph from the ASD dataset according to the architecture and dataset for pretraining. Grad-CAM visualized the area that the models focused on (red and yellow). All models gave the correct answer. CM; pretrained on the ChestX-ray dataset, IN; pretrained on the ImageNet dataset.

## Discussion

In this study, we developed deep learning-based predictive models for diagnosing ASD on chest radiographs using a method of transfer learning, and validated that the diagnostic performance of the models was comparable to that of expert cardiologists. ASD is characterized by an abnormal opening in the atrial septum that allows shunting of blood flow between the left and right atria, leading to an increase in pulmonary flow and enlargement of right heart structures [3, 4]. Accordingly, chest radiographic findings in ASD typically include increased caliber of pulmonary arteries with prominent pulmonary vasculature and enlargement of right atrium and ventricle with leftward cardiac rotation [3, 4]. Chest radiography is quickly and easily performed, and thus considered as the first step imaging test in general practice and primary care used to help diagnose cardiovascular and pulmonary diseases. In Japan, routine health check-ups are available to almost all segments of population throughout their life course [28]. Students and full-time employees, as well as people aged between 40 and 74, are legally required to have annual health check-ups, and other comprehensive health check-ups are also available from municipalities, insurers, and providers. This unique healthcare system provides relatively frequent opportunities for people to have chest radiographs while they are young and have no current health problems. However, physicians except for expert cardiologists or radiologists have difficulties in making accurate diagnosis of ASD based on chest radiographs alone, and the diagnostic value of chest radiography is limited, especially in early stage of ASD. Our deep-learning-based models on chest radiographs reached the comparable level to expert cardiologists in the accuracy of detecting ASD. Although further improvement in accuracy is needed, future implementation of our models in healthcare will offer a solution to improve clinical workflow and diagnostic performance, leading to reduction in the number of ASD patients who are otherwise overlooked and undiagnosed for decades.

ASD accounts for approximately 10 to 15 percent of congenital heart disease (CHD), with an estimated birth prevalence of approximately 1 to 2 per 1,000 live births [29]. In Japan, the number of adults with CHD were estimated to be 409,101 in 2007, with an annual increase of 9,000 [30]. Accordingly, there are currently at least 500,000 adults with CHD in Japan. Since ASD is reported to account for 25–30% of CHD cases diagnosed in adulthood [4], the prevalence of adult ASD is estimated at 1.5 per 1,000 in Japan. If we apply our model (e.g. ResNet50-based model pretrained on the ImageNet dataset; sensitivity 0.86 and specificity 0.87) to 100,000 Japanese adults, true positives/false positives/false negatives would be 129/12981/21, respectively. The false positives are large in number in our model, and we need to improve the performance of our models, or use other modalities together with our models to diminish the false positives.

Transfer learning is a useful method to address the problem of training a classifier when large and complete annotated datasets for training are not available [31]. For medical imaging, transfer learning is typically performed by taking the standard model designed for the large-scale annotated natural image dataset such as ImageNet [19] with corresponding pretrained weights, and then fine-tuning the model on the limited medical imaging dataset [32, 33]. However, there exist inherent differences in data sizes, features, and task specifications between natural and medical imaging datasets, and precise effects of the natural image dataset on performance gains are undetermined [34, 35]. We therefore hypothesized that pretraining on a dataset with similar task specification (e.g. interpretation of chest radiographs) would improve the performance, and selected the ChestX-ray14 chest X-ray dataset for pretraining of another model. Although both models were able to detect chest radiographic abnormalities associated with ASD with high accuracy, Grad-CAM showed that these models focused on different areas in the chest radiographs. It is currently unclear why the areas that the models considered important for detecting ASD are different, because the basis for making a judgment by a deep learning-based algorithm is incomprehensible inside “black box”. These findings also indicate that ensemble learning by combining these models will help to obtain better performance for detecting ASD [36].

There are several limitations in our “proof-of-concept” study. First, this is a single-center study, and further study is needed using a larger dataset including external data. Second, many patients with severe ASD were included in our dataset, who had high pulmonary to systemic flow ratio or pulmonary hypertension, and the chest radiographic findings in these patients might be relatively apparent. Third, the control group in our study included patients who had coronary angiography or percutaneous coronary intervention, because they were confirmed not to have ASD by transthoracic echocardiography, and their chest radiographs assumably represented those of general population. Ideally, the control group should consist of healthy subjects for the purpose of implementing our models to screen ASD on chest radiographs in primary care or health check-ups. Fourth, other congenital heart diseases and cardiac diseases with pulmonary hypertension were excluded in this study, and our models might not distinguish images of ASD patients from those of patients with these cardiac diseases.

The present study demonstrated that deep learning-based models could detect abnormal signs in chest radiographs suggesting ASD with high accuracy equivalent to expert cardiologists. Further studies using a larger and broader dataset will improve the diagnostic performance of the models, and determine whether deep learning-based approach is feasible to help clinicians make accurate diagnosis of ASD in clinical setting.

## Data Availability

All data produced in the present study are available upon reasonable request to the authors.

## Supporting information

**S1 Fig. Performance detecting cardiomegaly of the model pretrained on the ChestX-ray dataset before fine-tuning**.

A. ROC curves for the models on the test set of the ChestX-ray dataset just after pretrained on the ChestX-ray dataset.

B. Chest radiographs in the test set of the ChestX-ray dataset, which was labelled with “Cardiomegaly”. Grad-CAM shows the area that the model focused on.

## Notes

### Competing Interest Statement

The authors have declared no competing interest.

### Funding Statement

This study did not receive any funding.

### Author Declarations

The Institutional Review Board of the University of Tokyo gave ethical approval for this work.

