## Supplementary material for "Deep learning-based approach for detecting signs of atrial septal defect on chest radiographs: a proof of concept study": S1 Fig

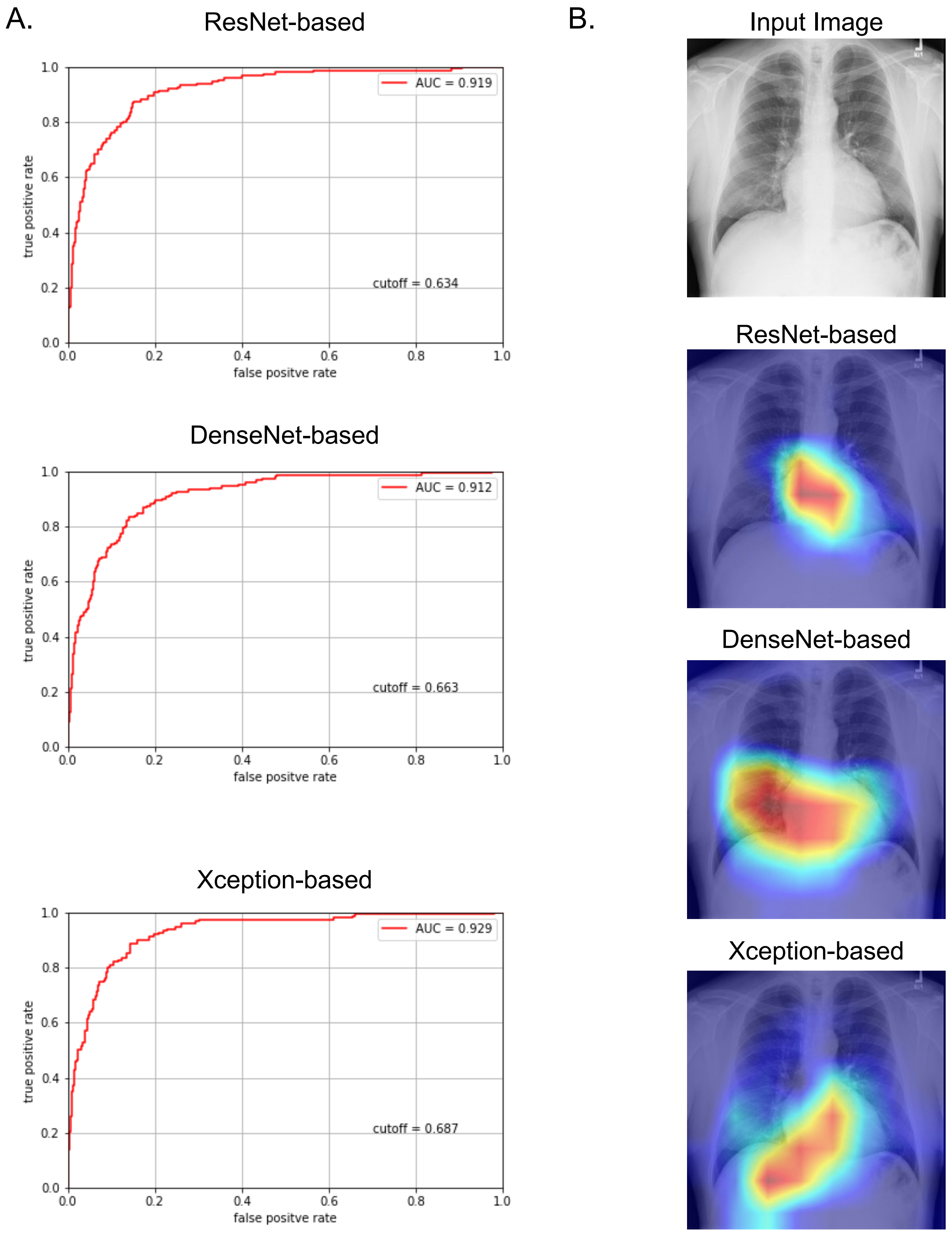


**S1 Fig. Performance detecting cardiomegaly of the models pretrained on the ChestX-ray dataset before fine-tuning.**

A. ROC curves for the models on the test set of ChestX-ray dataset just after pretrained on the ChestX-ray dataset.

B. Chest radiographs in the test set of the ChestX-ray dataset, which was labelled with “Cardiomegaly”. Grad-CAM shows the area that the model focused on.
